# The value of angiogenic biomarkers in determining preeclampsia-related perinatal deaths: a cross-sectional study in a high burden setting of southern Mozambique

**DOI:** 10.64898/2025.12.15.25342319

**Authors:** Maureen N. Chileshe, Natalia Rakislova, Carla Carrilho, Fabiola Fernandes, Tacilta Nhampossa, Alba Morató, Lorena Marimon, Núria Peñuelas, Anete Mendes, Elvira Luis, Jahit Sacarlal, Isaac Casas, Jessica Navero-Castillejos, Antía Figueroa-Romero, Manuel Morales-Ruiz, Juan Carlos Hurtado, Miguel J. Martinez, Jaume Ordi, Clara Menendez, Raquel González

## Abstract

**Background:** Estimating the contribution of preeclampsia (PE) to perinatal mortality in low-resource settings remains difficult due to limited diagnostic capacity. Lack of specific histopathologic changes in stillbirth, foetuses and/or placenta often hampers assignment of PE as cause of death. While PE angiogenic biomarkers (sFlt-1, PlGF) predict adverse pregnancy outcomes, their post-mortem diagnostic utility is unclear. We evaluated the potential role of these biomarkers in identifying PE-related perinatal deaths.

**Methods:** This cross-sectional study was conducted among pregnant women who delivered a stillborn infant or experienced an early neonatal death at Maputo Central Hospital in Mozambique. Concentrations of sFlt-1, PlGF and their ratio (sFlt-1/PlGF) were assessed in maternal blood and post-mortem foetal/neonatal and placental blood. Causes of death were determined via minimally invasive tissue sampling (MITS) and diagnostic accuracy was assessed using ROC curves for standardized and optimal biomarker cut-offs.

**Results:** A total of 100 women with perinatal deaths (98 stillbirths and 2 early neonatal deaths) were included in the study between March 2021 and April 2022. Maternal sFlt-1/PlGF ratios showed the highest diagnostic accuracy for PE-related deaths (area under curve [AUC]=82%), followed by placental (AUC=75%) and foetal blood (AUC=61%). PlGF levels were significantly reduced in PE-associated deaths. sFlt-1/PlGF cutoffs of ≥85 and ≥110 in maternal and placental blood were reliable cut-offs for identifying PE-related foetal/neonatal deaths. Optimal ratio cut-offs were ≥50 (maternal blood), ≥230 (placental blood), and ≥140 (foetal blood), with maternal and placental sFlt-1/PlGF ratios showing significant associations with PE-related death (OR=10.58 and 5.98 respectively). Sensitivity and specificity in maternal and placental blood were 84% and 73%, and 67% and 78%, respectively, with a positive predictive value (PPV) of 84% in both.

**Conclusions:** Maternal and placental angiogenic biomarkers enable reliable identification of preeclampsia-related perinatal deaths and could significantly enhance cause-of-death determination in low-resource, high-burden settings. These findings underscore the potential value of biomarker measurement for both risk stratification and mortality surveillance.

**Research in context:** *What is already known on this topic:*

- sFlt-1, PlGF, and their ratio, are well-established in predicting and diagnosing preeclampsia during pregnancy.
- However, their utility in identifying preeclampsia-related perinatal deaths particularly in postmortem investigations such as minimally invasive tissue sampling (MITS) samples, is underexplored.

*What this study adds:*

- This study is the first to evaluate diagnostic accuracy of these biomarkers using postmortem placental and foetal/neonatal blood samples from MITS to identify preeclampsia-related perinatal deaths.
- We show that maternal and placental sFlt-1/PlGF ratios can reliably identify preeclampsia-related perinatal deaths at standardized ratio cutoffs of ≥85 and ≥110 respectively. Whilst significantly reduced PlGF concentrations, lower placental and perinatal anthropometric measurements were hallmark features of PE-related deaths.
- The study suggests optimal sFlt-1/PlGF ratio cut-offs for maternal (≥50) and placental (≥230) blood, providing robust diagnostic thresholds for use in perinatal mortality surveillance and antenatal risk stratification in high-burden settings.

*How this study might affect research, practice or policy:*

- The use of preeclampsia biomarker analysis in determining causes of perinatal death offers a feasible, scalable, and objective method that may help overcome challenges in accurate cause of death attribution in LMICs.
- This has important implications for mortality surveillance system monitoring and pregnancy risk assessments, where autopsy is rare and clinical records are often incomplete.
- Incorporating biomarker-based diagnostics into existing perinatal death investigations could significantly improve mortality data and guide targeted health interventions.

## Introduction

Global under-five mortality has declined by 51% since 2000, yet perinatal deaths remain a persistent challenge, accounting for nearly half of child mortality alongside 1.9 million annual stillbirths.^1^ The burden is disproportionately concentrated in low- and middle-income countries (LMICs), with sub-Saharan Africa and South Asia collectively accounting for 75% of cases.^1^ Hypertensive disorders of pregnancy, particularly preeclampsia (PE), are estimated to contribute substantially to this mortality burden.^2,3^ However, accurate attribution of deaths to PE remains problematic in resource-constrained contexts due to high rates of non-institutional deliveries and limited diagnostic tools.^3,4^

PE is a multisystem hypertensive disorder affecting 2-8% of pregnancies characterized by new onset hypertension after 20 weeks of gestation associated with proteinuria and/or end organ dysfunction.^5^ Abnormal placentation and impaired placental angiogenesis leading to reduced placental-foetal blood flow are believed to contribute to the development and progression of PE.^6,7^ This disruption results in an abnormal regulation of angiogenic factors, evident with the elevated soluble fms-like tyrosine kinase-1 (sFlt-1) and reduced placental growth factor (PlGF), which triggers systemic vascular dysfunction and the associated clinical manifestations and impairs foetal nourishment and development resulting in adverse perinatal outcomes.^5,8^

The clinical utility of these angiogenic biomarkers in PE prediction is well-established.^6–9^ Multicentre studies have shown that an sFlt-1/PlGF ratio <38 reliably excludes PE development within one week, while low thresholds of PlGF in pregnant women was associated with a higher rate of perinatal deaths and growth restriction.^10,11^ Elevated ratios are also strongly linked with poor foetal growth and stillbirth. ^6,7,10,11^

When reliable clinical histories are unavailable, attribution of stillbirth or early neonatal death to PE can be indirectly suggested by autopsy findings,^12^ including placental pathology.^13,14^ However, definitive confirmation is challenging without antenatal and obstetrical clinical data. Post-mortem angiogenic biomarkers (from foetal, neonatal, or placental blood) could be of diagnostic value in PE attribution, yet their utility remains underexplored. Our study therefore sought: (1) to assess the performance of sFlt-1, PlGF, and their ratio to identify PE-related perinatal deaths using maternal blood and post-mortem samples i.e foetal/neonatal and placental blood; (2) to estimate and compare biomarker concentrations in foetal/neonatal and placental blood with their levels in maternal blood of women with and without perinatal loss; and (3) to correlate perinatal anthropometry and placental measurements with PE-related deaths.

## Methods

### Study setting and design

This cross-sectional study was conducted at the Maputo Central Hospital (MCH), a referral quaternary hospital in Mozambique situated in the capital city, Maputo. The hospital serves a population of three million inhabitants with an estimated 400–500 perinatal deaths annually. The average stillbirth rate in the country is 17 per 1,000 total births, and 41% of the under-five deaths are neonatal deaths. ^15^ Estimated neonatal mortality rate in Maputo city and province in 2020 was 10 and 14 deaths per 1,000 live births, respectively.^16^ Additionally, data from an autopsy study on causes of perinatal death carried out at this institution showed that PE contributed to foetal mortality in 25% to 30% of stillbirths.^17^

### Study procedures and sample collection

The study protocol and informed consent form were reviewed and approved by the National Committee on Health Bioethics of Mozambique (Ref: 392/CNBS/20). All methods were carried out in accordance with relevant guidelines and regulations, including the last version of the Declaration of Helsinki.

Women delivering a stillborn or having a very early neonatal death (death occurring within the first 12 hours of life) at the MCH, were invited to take part in the study. After explaining the study procedures, eligible participants who gave written informed consent were enrolled. Five mL of maternal venous blood were collected for the analysis of angiogenic biomarkers. Demographic, clinical and obstetric/antenatal characteristics of the mothers, including delivery information were recorded in a REDCap-designed electronic database.

Sample collection procedures were conducted between 1 March 2021 and 1 April 2022. Blood samples were collected for angiogenic biomarkers analysis from all perinatal deaths and placentas during the minimally invasive tissue sampling (MITS) procedure: 20 mL of foetal/neonatal blood extracted by puncture of the subclavian vein,^18,19^ and 5 mL of placental blood collected by puncture from the maternal side with a syringe. Additionally, available samples from 42 pregnant women who delivered live births were used as “controls” for the analysis of the levels of angiogenic biomarkers. Women providing these control samples were participating in a malaria prevention clinical trial during the same study period in a neighboring study site.^20^ Informed consent was obtained from all the participants and their data collected in an OpenClinica clinical trial data base.^20^

MITS was performed using a simplified protocol 6-24 hours after demise.^18,19^ The protocol also included evaluation of the foetus and the placenta. The following anthropometric measures were recorded: foetal weight, length, head and abdominal circumferences. Any external anomalies including malformations, skin lesions and maceration were registered. Tissue sampling for microbiology from both sides of the thorax (lungs and heart) using a 16G biopsy needle (Bard Monopty), followed by collection of samples from both sides of the thorax for histological evaluation. The placenta was photographed, measured and weighed. Samples of placental tissue from the maternal and foetal sides, and the umbilical cord were collected for microbiology. The placenta was then fixed *in toto* in 10% neutral buffered formalin for 24-72 hours and processed following the Amsterdam placental workshop consensus guidelines.^21^

### Laboratory procedures and cause of death determination

Histopathological, biochemical and microbiological analyses on the collected samples were performed at the Hospital Clínic of Barcelona (Spain), as explained in detail elsewhere.^18,19^ Concentrations of sFlt-1 and PlGF biomarkers were measured in maternal, foetal/neonatal and placental sera using an automated Elecsys® assay kit on an electrochemiluminescence platform Cobas 6000 analyser system (Roche Diagnostics, Penzberg, Germany). These values were then used to calculate the sFlt-1/PlGF ratio. The intra- and inter-assay coefficients of variation for both biomarkers were lower than 4% and 8%, respectively. For the prediction of PE-related perinatal deaths, standardized cut-off values of sFlt-1/PlGF ratio (≥38, ≥85 and ≥110) were used. ^22^ In addition, optimal cut-offs of the sFlt-1/PlGF ratio that could identify PE-related perinatal deaths in the study group were calculated.

A panel composed of a pathologist, an obstetrician and a microbiologist, evaluated the clinical antenatal and obstetrical information, as well as the MITS report, including gross (photographs, measurements), microbiological and histological post-mortem evaluation of the foetus and placenta. The members of the panel were blind to the angiogenic biomarker results.^18,19^ Main (foetal and maternal) causes of death, as well as contributing causes to death conditions were assigned using the World Health Organization international classification of diseases-10 to deaths during the perinatal period (ICD-PM).^17^

### Definitions and diagnostic criteria

PE was defined as the presence of de novo hypertension (systolic blood pressure ≥140 mmHg and/or diastolic pressure ≥90 mmHg) after 20 weeks’ gestation accompanied by proteinuria and/or evidence of maternal acute kidney injury, liver dysfunction, neurological features, hemolysis or thrombocytopenia, and/or foetal growth restriction.^5^ Maternal PE comprised of women with confirmed PE or eclampsia. A diagnosis of PE-related perinatal death (primary outcome) was defined as a death in which PE or eclampsia were identified by the panel as the main or contributing maternal condition leading to the death of the foetus or newborn. We classified newborns who died within the first 12 hours of life as very early neonatal deaths. Optimal cutoff values of the sFlt-1/PlGF ratio were cut-points that correctly classified the participants into having or not having a PE-related perinatal death.

### Statistical analysis

Statistical analysis was conducted using R Statistical Software (version 4.3.2; R Core Team, 2024). Categorical variables were presented as absolute frequencies and percentages, with comparisons made using the Chi-square or Fisher’s exact test.

Quantitative variables were described using medians and interquartile ranges (IQR), and comparisons between groups were performed using the Wilcoxon test.

Performance of different sFlt-1/PlGF ratio cut-offs in identifying PE-related perinatal deaths was assessed using sensitivity, specificity, accuracy, positive, and negative predictive value (PPV, NPV). Receiver operating characteristic (ROC) curve analysis was applied to determine the diagnostic accuracy of sFlt-1/PlGF ratio in each type of blood sample by the area under the curve (AUC) and optimal cut-points. Optimal cut-off values were defined as those that maximised the Youden index (difference between true positive and false positive rates). Univariate and multivariate logistic regression models were used to identify factors associated with PE-related perinatal deaths. Multivariable logistic regression models were built per blood sample by including all variables with a p-value<0.25 in the univariate analyses, followed by forward-backward stepwise selection adjusted by gestational age. A lower Akaike Information Criterion (AIC) was used to identify the best fit model.

## Results

### Baseline characteristics of participants

A total of 100 women who delivered a stillbirth or a baby who died within 12 hours after birth were included in the study from March 2021 to April 2022. Demographic and clinical features of the participants and their offspring are presented in table 1. The mean maternal age of study participants was 29.4 years (standard deviation, ±6.7), and 62 (62%) of them were multigravidas with a median gestational age of 34 weeks [IQR: 31, 37]. Mean systolic and diastolic arterial blood pressure at admission were 147 ± 31 mmHg, and 98 ± 20.3 mmHg. Ninety-eight (98.0%) babies were stillborn and two (2.0%) were very early neonatal deaths who were born with an Apgar score at 5-mins ≤ 3. The majority (72.0%) of the cases were preterm with low birth weight (1680 g; IQR: 1182.5, 2415.5). Of 98 stillbirths, 70 (71.4%) showed maceration at the MITS: 19 (27.2%) had grade 1, 15 (21.4%) grade 2, and 36 (51.4%) grade 3 maceration.

**Table 1.** Maternal, perinatal and placental characteristics of study participants.

|  | Women with perinatal deaths<br>(N = 100) |
| --- | --- |
| **Maternal Characteristics** |  |
| Maternal age | 29.4 ($\pm 6.7$)<sup>1</sup> |
| Gravidity |  |
| 1 | 23 (23.0%)<sup>2</sup> |
| 2-4 | 62 (62.0%)<sup>2</sup> |
| $\geq 5$ | 15 (15.0%)<sup>2</sup> |
| Gestational age at delivery (weeks) | 34.2 (31.1, 37.2)<sup>3</sup> |
| Type of delivery |  |
| Vaginal | 89 (89.0%)<sup>2</sup> |
| Caesarean section | 11 (11.0%)<sup>2</sup> |
| Severe anaemia (Hb<7g/dL) | 4 (5.2%)<sup>2</sup> |
| Missing information | 23 |
| Syphilis | 1 (1.1%)<sup>2</sup> |
| Missing information | 10 |
| HIV | 16 (16.0%)<sup>2</sup> |
| Systolic blood pressure (mmHg) | 138.5 (124.5, 162.0)<sup>3</sup> |
| Diastolic blood pressure (mmHg) | 95.0 (82.5, 107.5)<sup>3</sup> |
| Preeclampsia | 67 (67.0%)<sup>2</sup> |
| **Perinatal Characteristics** |  |
| Gestational Age at birth (weeks) |  |
| 20-27 | 3 (3.0%)<sup>2</sup> |
| 28-36 | 69 (69.0%)<sup>2</sup> |
| $\geq 37$ | 28 (28.0%)<sup>2</sup> |
| Sex |  |
| Female | 35 (35.0%)<sup>2</sup> |
| *Male* | 65 (65.0%) <sup>2</sup> |
| Birth weight (grams) | 1680.0 (1180.0, 2445.0) <sup>3</sup> |
| Height/length of the baby (cm) | 42.0 (39.0, 47.0) <sup>3</sup> |
| Head circumference of the baby (cm) | 29.5 (26.0, 32.0) <sup>3</sup> |
| Preeclampsia as main or contributing maternal cause of perinatal death (as attributed by cause of death panel) | 63 (63.0%) <sup>2</sup> |

<sup>1</sup> Mean (standard deviation); <sup>2</sup> absolute number (percentage); <sup>3</sup> median (interquartile range)

### Characteristics of the participants according to PE-related perinatal death status

Maternal characteristics, perinatal and placental anthropometric measurements were compared between participants with and without PE-related foetal/neonatal death (Supplementary Appendix, Table S1). Mean systolic and diastolic arterial blood pressure was 161.5 ± 29.6 and 121.6 ± 11.6 mmHg in women with PE-related perinatal deaths and 108.1 ± 17.8 and 80.1 ± 9.0 mmHg in women with perinatal deaths not associated with PE (p<0.001). Stillbirths/neonates whose death was related to maternal PE had a low median gestational age at birth (33 weeks), lower birth weight (1450 g), smaller body length (41 cm) and head circumference (28 cm) compared to those whose deaths were not related to PE. Similar differences were observed in placental measurements, with the PE-related perinatal death group showing lower mean placental weight, disc length/width, and umbilical cord diameter compared to the non-PE-related perinatal death group. Maternal PE was present in 67 women (67.0%), and was assigned the main cause or contributing factor of death in 50 and 13 babies, respectively, accounting for 63 (63.0%) of all perinatal deaths.

Among 98 stillbirths (Table S2), the main foetal causes of death were intrauterine hypoxia (71.4%, n=70), predominantly associated with maternal hypertensive disorders (preeclampsia/eclampsia) and placental/cord complications (Table S3); and pneumonia (24.5%, n=24). Severe birth asphyxia (Apgar scores 1 and 2 in the first minute of life) was the main cause of neonatal death.

### PE-angiogenic biomarker concentrations and diagnostic performance

Table 2 shows the biomarkers concentrations (sFlt-1, PlGF, and sFlt-1/PlGF ratio) in maternal, placental and foetal/neonatal blood in the perinatal death group and in maternal samples of the 42 controls. The perinatal death group showed significantly elevated median maternal sFlt-1 levels compared to controls (7788.5 pg/mL vs 5576.5 pg/mL, p=0.010) and markedly lower PlGF (74.6 pg/mL vs 154.5 pg/mL, p=0.039). Notably, women with PE-related perinatal deaths had sevenfold lower median PlGF than non-PE-related deaths (47.3 pg/mL vs 332.0 pg/mL, p<0.001). Overall, maternal sFlt-1/PlGF ratio was 3.3 times higher in women with perinatal deaths (107.4) than those with live births (32.8) (p<0.001), with an 8.1 times higher threshold in women with perinatal deaths linked to PE than non-PE deaths (21.30 vs 171.98, p<0.001). Placental blood showed similar trends of significantly higher sFlt-1/PlGF ratios in PE-related deaths than non-PE-related deaths (284 vs 110, p<0.001), whilst foetal/neonatal blood showed non-significant trends. In 76.2% (48/63) of PE-related perinatal deaths, the maternal sFlt-1/PlGF cut-off was ≥85, and 85.7% (54/63) with placental ratio cut-off ≥110 (Table S4). Women with PE who had live births or non-PE-related deaths showed lower sFlt-1/PlGF thresholds (Figure S1).

**Table 2.** Angiogenic biomarker concentrations in maternal, foetal/neonatal and placental blood.

|  | Perinatal death group<br>(N = 100) | Women with live birth<br>(controls),<br>(N = 42) | p-value<sup>1</sup> | Perinatal death group |  | p-value<sup>1</sup> |
| --- | --- | --- | --- | --- | --- | --- |
|  |  |  |  | No preeclampsia-related perinatal<br>death (N = 37) | Preeclampsia- related<br>perinatal death<br>(N = 63) |  |
| Maternal blood |  |  |  |  |  |  |
| sFlt-1 | 7788.5<br>(4445.5, 12366.0) | 5576.5<br>(3647.0, 7341.0) | 0.010 | 5854.0<br>(2595.0, 11787.0) | 8551<br>(4687.0, 13333.0) | 0.068 |
| PIGF | 74.6<br>(34.0, 281.5) | 154.5<br>(84.2, 244.0) | 0.039 | 332<br>(109.0, 683.0) | 47.3<br>(29.0, 110.0) | <0.001 |
| sFlt-1/PIGF ratio | 107.4<br>(24.0, 248.7) | 32.8<br>(15.3, 66.9) | <0.001 | 21.13<br>(10.4, 65.2) | 172.0<br>(92.7, 392.3) | <0.001 |
| Placental blood |  |  |  |  |  |  |
| sFlt-1 | 440770.0<br>(188410.0, 739355.0) |  |  | 363750.0<br>(111570.0, 653090.0) | 487650.0<br>(226080.0, 834330.0) | 0.11 |
| PIGF | 2042.5<br>(1091.0, 4555.5) |  |  | 3417.0<br>(1228.0, 6592.0) | 1835.0<br>(862.0, 3389.0) | 0.036 |
| sFlt-1/PIGF ratio | 234.7<br>(104.7, 385.5) |  |  | 110.2<br>(64.1, 216.6) | 284.9<br>(149.6, 420.8) | <0.001 |
| Foetal/neonatal blood |  |  |  |  |  |  |
| sFlt-1 | 28187.0<br>(11681.5, 114270.0) |  |  | 19777.0<br>(9563.0, 72550.0) | 36390.0<br>(15707.0, 115580.0) | 0.12 |
| PIGF | 239.5<br>(114.9, 617.5) |  |  | 282.0<br>(161.0, 580.0) | 215.0<br>(98.6, 655.0) | 0.3 |
| sFlt-1/PIGF ratio | 140.5<br>(70.7, 333.3) |  |  | 103.45<br>(43.9, 279.2) | 170.7<br>(86.3, 369.9) | 0.080 |

Values shown are median and (interquartile range)
<sup>1</sup> Wilcoxon rank sum test

The performance of the sFlt-1/PlGF ratio in identifying PE-related perinatal mortality is shown in Figure 1 and Table 3. The sFlt-1/PlGF ratio showed optimal diagnostic performance for identifying PE-related perinatal deaths in maternal blood (AUC=82%) outperforming placental (AUC=75%) and foetal/neonatal blood (AUC=61%). In maternal blood, a cut-off ≥50 provided the highest predictive value (PPV 84%, specificity 73%, sensitivity 84%) in identifying PE as the main and contributing cause of death, while ≥38 offered nearly balanced sensitivity/specificity (86%/68%) and ≥85 maximized specificity (78%) with a PPV of 86% and sensitivity of 76%. Placental blood showed moderate diagnostic utility; a ratio cut-off of ≥110 yielded a sensitivity of 86%, specificity of 49%, a PPV of 74% with an accuracy of 72%; a ≥230 cutoff matched maternal ≥50 PPV (84%) with a sensitivity of 67%, specificity of 78%, a PPV of 84% and accuracy of 71%. Foetal/neonatal blood showed limited overall diagnostic value. Of note, sFlt-1/PLGF ratio cut-offs of ≥100 (maternal), ≥240 (placental) and ≥140 (foetal) were optimal values in identifying PE as the main cause of perinatal death (Figure S2).

**Figure 1.**
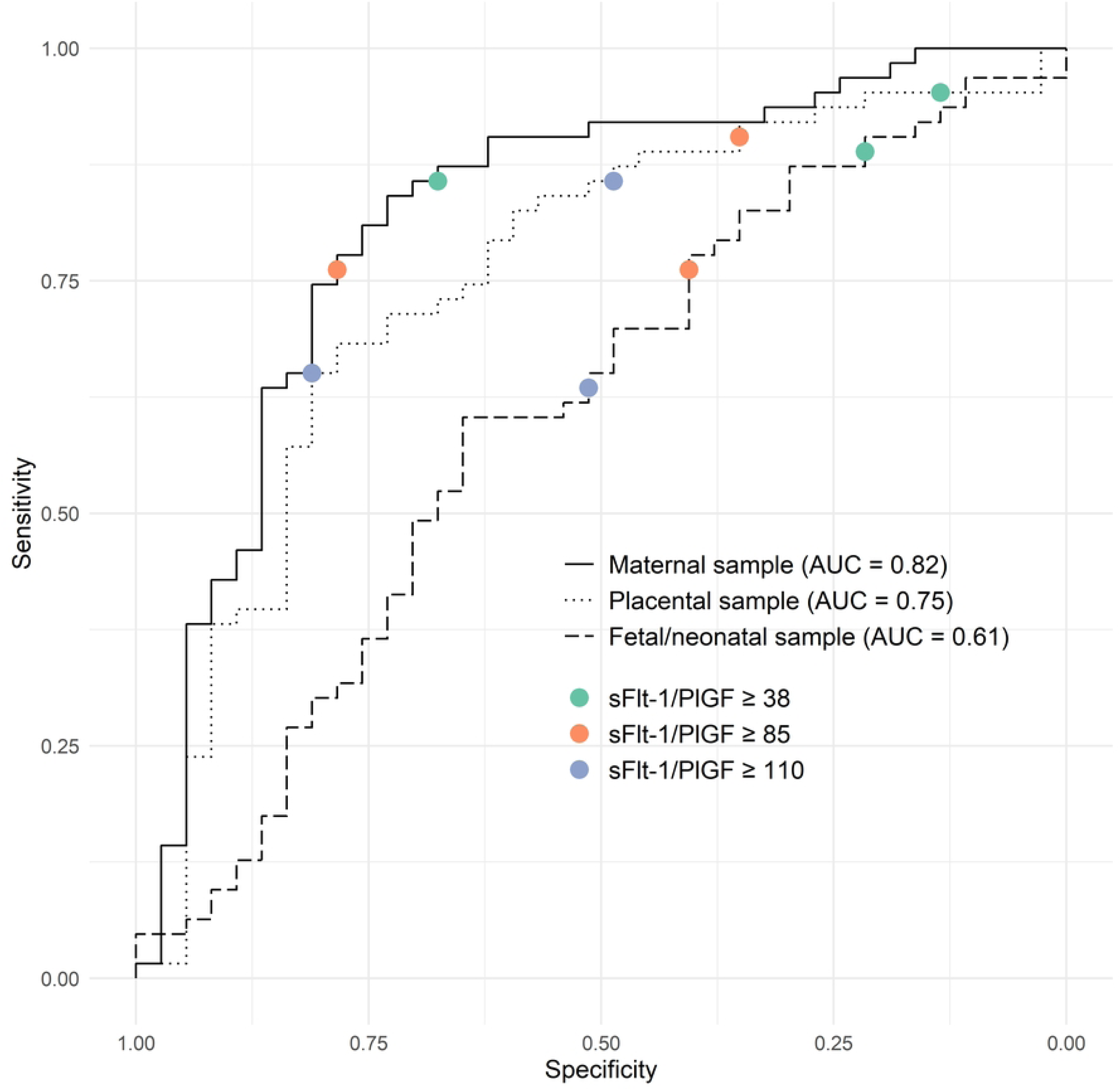
Factors associated with preeclampsia-related perinatal death using standardized biomarker cut-offs.

**Table 3.** Sensitivity, specificity, accuracy, positive predictive value (PPV), negative predictive value (NPV) values of sFlt-1/PlGF ratio at different cut-offs to identify preeclampsia related perinatal death.

| Blood sample | Sensitivity<sup>1</sup> | Specificity<sup>1</sup> | Accuracy<sup>1</sup> | PPV<sup>1</sup> | NPV<sup>1</sup> |
| --- | --- | --- | --- | --- | --- |
| **Standardized cut-offs** |  |  |  |  |  |
| **sFlt-1/PIGF $\geq 38$** |  |  |  |  |  |
| Maternal | 86 (75 - 93) | 68 (50 - 82) | 79 (70-87) | 82 (70-90) | 74 (56-87) |
| Placental | 95 (87 - 99) | 14 (05 - 29) | 65 (55-74) | 65 (55-75) | 62 (24-91) |
| Foetal/neonatal | 89 (78 - 95) | 22 (10 - 38) | 64 (54-73) | 66 (55-76) | 53 (27-79) |
| **sFlt-1/PIGF $\geq 85$** |  |  |  |  |  |
| Maternal | 76 (64 - 86) | 78 (62 - 90) | 77 (68-85) | 86 (74-94) | 66 (50-80) |
| Placental | 90 (80 - 96) | 35 (20 - 53) | 70 (60-79) | 70 (59-80) | 68 (43-87) |
| Foetal/neonatal | 76 (64 - 86) | 41 (25 - 58) | 63 (53-72) | 69 (56-79) | 50 (31-69) |
| **sFlt-1/PIGF $\geq 110$** |  |  |  |  |  |
| Maternal | 65 (52 - 77) | 81 (65 - 92) | 71 (61-80) | 85 (72-94) | 58 (43-71) |
| Placental | 86 (75 - 93) | 49 (32 - 66) | 72 (62-81) | 74 (62-84) | 67 (46-84) |
| Foetal/neonatal | 64 (50 - 75) | 51 (34 - 68) | 59 (49-69) | 69 (56-81) | 45 (30-61) |

| Optimal cut-offs |  |  |  |  |  |
| --- | --- | --- | --- | --- | --- |
| **sFlt-1/PIGF $\geq 50$** |  |  |  |  |  |
| Maternal | 84 (73 - 92) | 73 (56 - 86) | 80 (71-87) | 84 (73-92) | 73 (56-86) |
| **sFlt-1/PIGF $\geq 230$** |  |  |  |  |  |
| Placental | 67 (54 - 78) | 78 (62 - 90) | 71 (61-80) | 84 (71-93) | 58 (43-72) |
| **sFlt-1/PIGF $\geq 140$** |  |  |  |  |  |
| Foetal/neonatal | 59 (46 - 71) | 65 (47 - 80) | 61 (51-71) | 74 (60-85) | 48 (34-63) |
| Values shown are percentage and 95% confidence intervals |  |  |  |  |  |
| sFlt-1 - soluble fms-like tyrosine kinase-1, PIGF - Placental Growth Factor |  |  |  |  |  |

### Associations between biomarker thresholds, perinatal features and PE-related mortality

Figure 2 presents the sFlt-1/PlGF ratio cut-off values, perinatal features, and placental characteristics associated with PE-related perinatal mortality. In the multivariate models built for each blood compartment, maternal blood showed the best model fit with a cut-off of ≥85 (AIC=96.5), followed by placental blood (≥110, AIC=101) and foetal/neonatal blood (≥85, AIC=111). Among the perinatal and placental anthropometric variables, birth weight, placental weight, and umbilical cord diameter were retained in the models, while other highly correlated variables were excluded.

**Figure 2.**
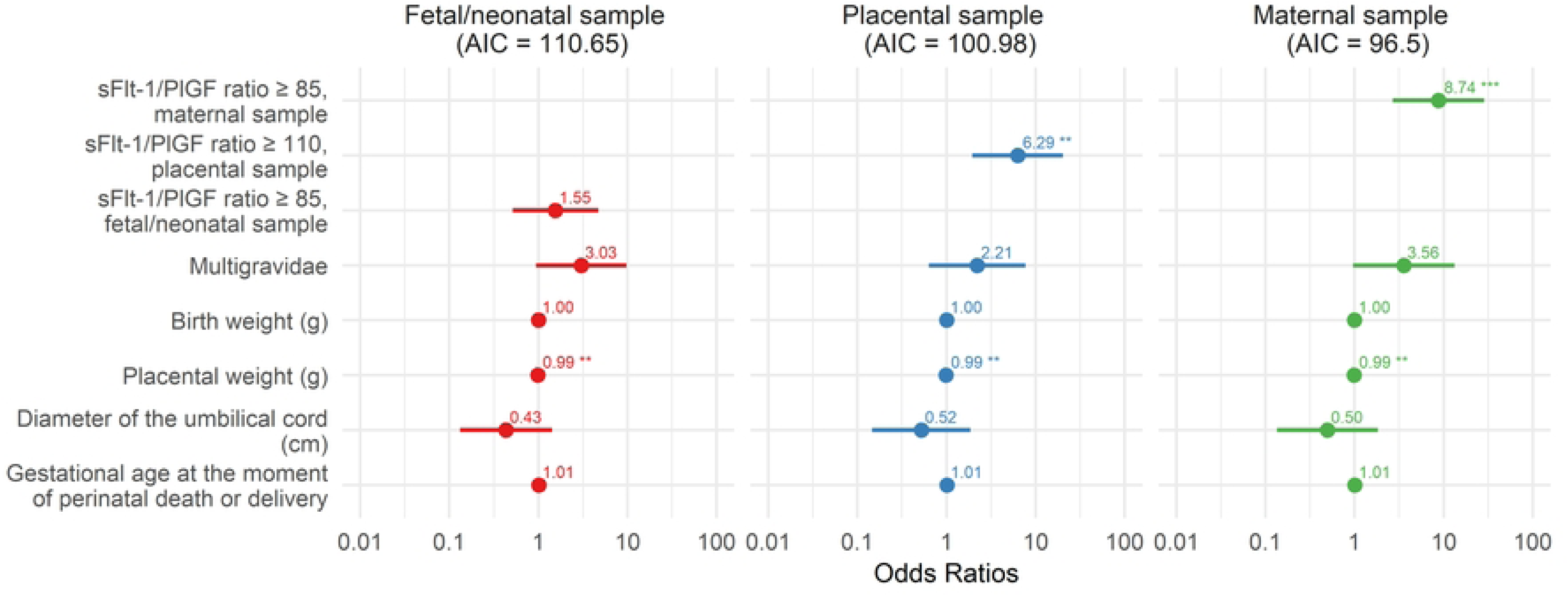
Factors associated with preeclampsia-related perinatal death using standardized biomarker cut-offs.

A maternal blood sFlt-1/PlGF ratio of ≥85 was significantly associated with an increased risk of PE-related perinatal death (OR 8.74; 95%CI: 2.82–30.81; p<0.001). Similarly, in placental blood, a ratio of ≥110 also showed a significant association (OR 6.29; 95%CI: 2.02–21.48; p=0.002). In contrast, foetal/neonatal blood (≥85) did not reach statistical significance (OR=1.55; 95%CI: 0.50–4.73; p=0.4). When considering previously identified optimal cut-offs of sFlt-1/PlGF ratio, cut-offs of ≥50 in maternal blood and ≥230 in placental blood (Figure 3), were both significantly associated with PE-related perinatal death (OR 10.58; 95%CI: 3.370–37.619; p<0.001 and OR 5.98; 95%CI: 2.029–19.924; p=0.002, respectively). Again, foetal/neonatal blood (≥140) did not show a significant association (OR=1.23, 95%CI: 0.42–4.46; p=0.7). Higher placental weight was associated with a reduced risk of having a PE-related perinatal death across all the models: maternal blood (OR 0.986; 95% CI, 0.976–0.995; p=0.003), placental blood (OR 0.986; 95%CI, 0.975–0.994; p=0.002), and foetal/neonatal blood (OR 0.988; 95% CI, 0.979–0.996; p=0.005).

**Figure 3.**
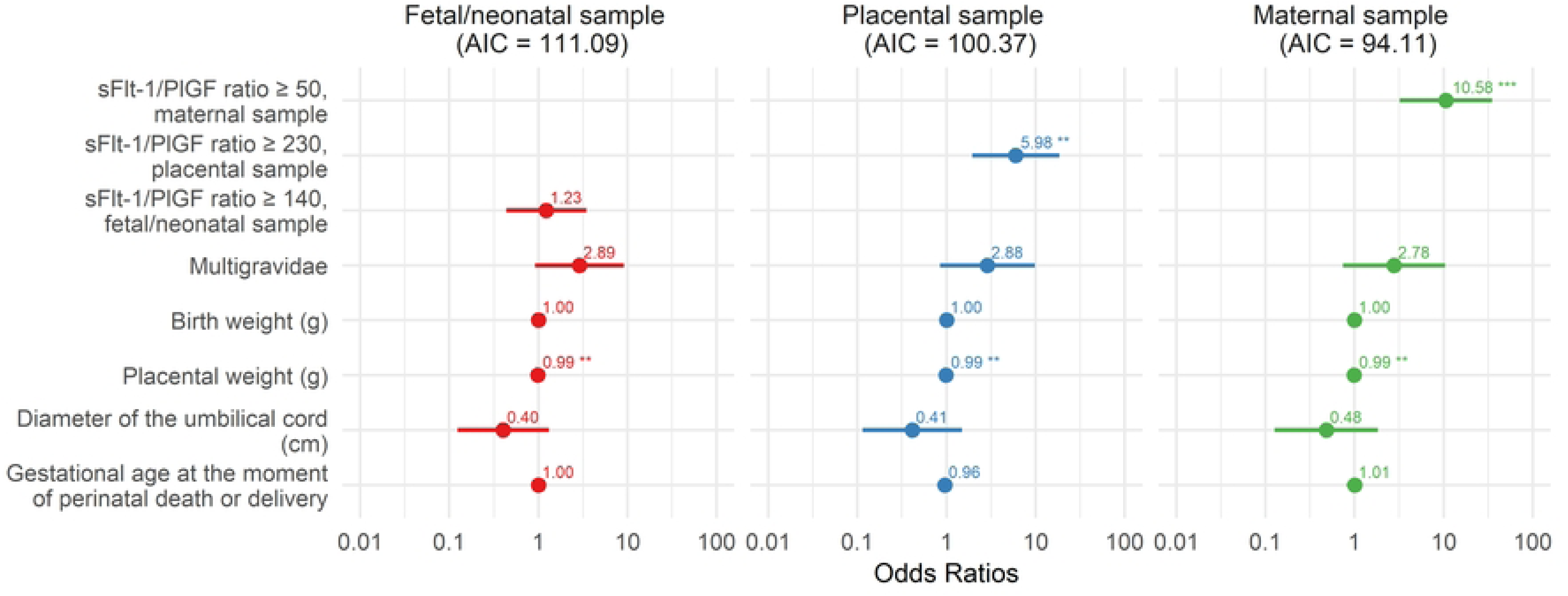
Factors associated with preeclampsia-related perinatal death using optimal biomarker cut-offs.

## Discussion

This study assessed the usefulness of sFlt-1, PlGF and their ratio in identifying PE-related perinatal deaths in Mozambican pregnant women with perinatal loss using maternal blood and post-mortem samples (foetal/neonatal and placental blood). To our knowledge, this is the first study evaluating angiogenic biomarkers in MITS blood samples for cause of death identification in stillbirths and early neonatal deaths. Maternal blood provided the highest diagnostic accuracy, followed by placental blood, whereas foetal/neonatal showed limited discriminatory capacity. We identified maternal sFlt-1/PlGF ratio cut-offs (≥85 standardized and ≥50 optimal) and placental ratio cut-offs (≥110 standardized and ≥230 optimal) as reliable thresholds in determining PE-related perinatal deaths and associated mortality risks in this population. Additionally, higher placental weight was associated with reduced risk of PE-related perinatal death – a supportive indicator for PE attribution.

The robust diagnostic performance of maternal sFlt-1, PlGF, and their ratio aligns with well-established evidence for predicting PE-related adverse outcomes. ^23–25^ A recent study from Germany found that 68% of 1117 pregnant women with adverse outcomes had a median sFlt-1/PlGF ratio of 177 (IQR: 54-362), with ≥85 threshold predicting preterm birth, respiratory distress, neonatal intensive care admission and stillbirth. ^26,27^ In another study, lower PlGF levels and higher sFlt-1/PlGF ratios were found in maternal blood of PE cases compared to non-PE cases (PlGF: 27.2 pg/mL vs 148.50 pg/mL, p<0.001; sFlt-1/PlGF: 378.5 vs 30.6, p<0.001, respectively. ^24^ Similar to our findings, low PlGF concentrations and elevated sFlt-1/PlGF ratios were associated with increased risk of having a perinatal death and severe complications. ^23–25^ Ratio cut-offs of ≥85 and ≥110 have been strongly linked to perinatal death, ^6–8,13,14,31,32^ with a report indicating that a sFlt-1/PlGF ratio of >85 predicts foetal compromise and preterm labour (PPV=86%, AUC=87%), especially before 34 weeks gestation in women with suspected/confirmed PE. ^28^ The strong diagnostic performance of these maternal blood biomarkers could enhance mortality surveillance, particularly in settings where autopsies are rarely conducted. On the other hand, an elevated (≥85 or ≥50) in high-risk pregnancies can play a crucial role in risk stratification, monitoring, and timely clinical intervention, potentially helping to reduce stillbirths.

Placental blood demonstrated acceptable diagnostic accuracy, with an optimal sFlt-1/PlGF cut-off (≥230) achieving an 84% PPV, similar to maternal blood. This supports its viability as an alternative diagnostic medium when maternal samples or clinical information are unavailable, especially in stillbirth investigations. ^29^ In line with placental dysfunction pathophysiology, PE-related deaths showed the triad of reduced birth weight, diminished placental weight, and narrower umbilical cord diameter. ^6,7,10,11,30,31^ The inverse association between placental weight and PE-related mortality further confirms placental insufficiency as a key driver of PE-related adverse outcomes. Even though birth weight was not statistically associated with PE-related perinatal deaths, the positive correlation between placental weight and birth weight is well established. ^11,27–29^ Both low birth weight and placental weight have been found to be associated with increased risk of placental malperfusion, foetal growth restriction and stillbirth/neonatal death. ^28,29^

Furthermore, our results show that intrauterine hypoxia was the leading cause of perinatal death, reinforcing this vicious cycle in which reduced placental-foetal blood flow leads to prolonged foetal hypoxia, triggering oxidative stress that further damages placental function and compromises foetal survival. ^32^ Placentas associated with stillbirths and foetal growth restriction in women with hypertensive disorders in pregnancy have a high incidence of vascular mal- or underperfusion, resulting in elevated OS biomarkers in foetal blood, making the foetus more susceptible to injury, prematurity and death. ^30–32^ These findings support integrating placental pathology with biomarker analysis in perinatal mortality investigations to improve cause of death attribution.

The limited diagnostic utility of foetal/neonatal sFlt-1/PlGF ratios for PE-related death could likely reflect a combination of factors including post-mortem degradation, variation of biomarker concentration due to collection site, type of blood sample (cord vs foetal blood) and the influence of gestational age or timing of death. While maternal and placental biomarker ratios showed consistent diagnostic performance, foetal measurements may have been compromised by autolytic changes, likely due to maceration, indicating the need for cautious interpretation in stillbirth cases. Future studies addressing these gaps including optimal sampling timing would be necessary in guiding biomarker analysis in post-mortem investigations.

The main strengths of this study are in its comprehensive multi-compartment sampling (maternal/foetal/placental), combined with placental and foetal anthropometrics data, and rigorous cause of death determination using WHO ICD-PM standards. The novel post-mortem application of angiogenic biomarkers and identification of reliable cut-offs in placental blood for PE-related mortality attribution offers a promising avenue for mortality surveillance when other information is unavailable. Furthermore, the demonstrated feasibility of MITS in a high-burden Mozambican population provides immediate relevant data for similar settings. The limitations include the cross-sectional design, which prevented us from tracking serial biomarker changes, a factor that could have improved outcome prediction and optimized the timing of testing. The high prevalence of foetal maceration may have compromised the pathological assessment of foetal tissues and foetal blood biomarker integrity. Therefore, larger longitudinal multi-center studies in high-burden settings are required to generate context-specific data for clinical guidelines and post-mortem protocols. With recent approval of biomarker immunoassays, ^33^ its implementation in resource-limited settings could additionally provide valuable evidence on feasibility and sustainability.

## Conclusion

In conclusion, angiogenic biomarkers in maternal and placental blood serve as reliable indicators of PE-related perinatal deaths. Post-mortem placental biomarkers can contribute to improving cause attribution to PE, an important advancement for high burden settings. Integrating biomarker testing into screening and post-mortem evaluations has the potential to accelerate the prevention of stillbirths and neonatal deaths while enhancing the accuracy of cause-of-death information.

## Data Availability

All relevant data are within the manuscript and its Supporting Information files.

## List of abbreviations

IC: Akaike Information Criterion
AUC: Area Under the Curve
CI: Confidence Interval
ICD-PM: International Classification of Diseases for Perinatal Mortality
IQR: Interquartile Range
LMICs: Low- and Middle-Income Countries
MCH: Maputo Central Hospital
MITS: Minimally Invasive Tissue Sampling
NPV: Negative Predictive Value
OR: Odds Ratio
PE: Preeclampsia
PlGF: Placental Growth Factor
PPV: Positive Predictive Value
REDCap: Research Electronic Data Capture
ROC: Receiver Operating Characteristic
sFlt-1: Soluble fms-like Tyrosine Kinase-1
WHO: World Health Organization

## Ethics approval and consent to participate

Ethics approval: The study protocol and informed consent form were reviewed and approved by the National Committee on Health Bioethics of Mozambique (Ref: 392/CNBS/20).

Patient consent: Written informed consent was obtained from the patient’s next of kin. Participants were provided with detailed information about the study before giving their written consent to be enrolled.

## Consent for publication

Not applicable

## Contributors

The study was conceived and designed by CM, JO and NR. TN, CC, AM, EL, JS, MM-R, MMa, MJM, RG and CM provided inputs on study design. Contributions to data acquisition were by TN, CC, AM, EL, JS, JNC, MM-R, MMa, MJM, JO, NR, RG, NP and CM. AM and MNC carried out data analysis and interpretation. The first draft of the manuscript was written by MNC and CM. RG and CM contributed equally to data interpretation and manuscript writing. All authors had access to the study data and provided critical revisions to the manuscript. All the authors hold the final responsibility for the decision to submit the manuscript for publication.

## Patient and public involvement

Neither patients nor the public were involved in the design, conduct, reporting, or dissemination plans of the research.

## Availability of data and materials

De-identified individual participant data (including biomarker values, cause-of-death classifications, and key clinical variables) that support the findings of this study are not publicly available due to sensitive nature of the study but are available from corresponding author on reasonable request.

## Acknowledgments

We extend our sincere gratitude to all study participants for their time and contributions, as well as to the research teams for their dedication to study procedures, technical support and coordination. We would also like to thank all the MIBio study personnel, Laura Garcia Otero and Mireia Piqueras for contributing to project coordination, and Sergi Sanz for his valuable support with statistical analysis and interpretation. We acknowledge support from the grant CEX2023-0001290-S funded by MCIN/AEI/10.13039/501100011033, and support from the Generalitat de Catalunya through the CERCA Program, the Department of Research and Universities of the Government of Catalonia (2021 SGR 01573); and CISM, which is supported by the Government of Mozambique and the Spanish Agency for International Development (AECID), for their support to the present publication.

